# Performance Analysis of Speech Recognition Models in Automated Scoring of the QuickSIN Test

**DOI:** 10.1101/2025.07.25.25332211

**Authors:** Arman Hassanpour, Yan Jiang, Paula Folkeard, Ewan Macpherson, Susan D. Scollie, Vijay Parsa

## Abstract

**Purpose:** Best practices in audiology recommend assessing speech understanding in noisy environments, especially for those with communication difficulties. Speech-in-noise (SiN) assessments such as the QuickSIN are used for validating signal processing in hearing aids (HAs) and are linked to HA satisfaction. This project seeks to enhance QuickSIN test efficiency by applying recent advancements in automatic speech recognition (ASR) technologies.

**Method:** Twenty-three adults with sensorineural hearing loss were fitted bilaterally with Unitron Moxi HAs and were administered the QuickSIN test in low and high reverberation environments. Testing was performed with two different HA programs: an omnidirectional program and a fixed directional microphone program. QuickSIN sentences were presented from 0° azimuth and competing babble from either 0°, laterally from 90° or 270°, or simultaneously from 90°, 180°, and 270° azimuths. Participants’ verbal responses to QuickSIN stimuli were scored by an audiologist and were recorded in parallel for offline transcription and scoring by ASR models from Amazon, Microsoft, NVIDIA, and Picovoice. The ASR-derived QuickSIN scores were compared to the corresponding audiologist-derived scores.

**Results:** Repeated Measures ANOVA results revealed that all ASR models overestimated the QuickSIN scores across most test conditions. Bland-Altman analyses showed that the Amazon ASR model had the least bias and the narrowest range for the limits of agreement, in comparison to the manual scoring by an experienced audiologist.

**Conclusions:** Some ASR models, such as Amazon, demonstrated performance comparable to that of an audiologist in automatically scoring QuickSIN tests. However, further refinements are necessary to increase the robustness of the ASR models in scoring low SNR loss test conditions.

## Introduction

Traditional audiology testing, which focuses on measuring pure-tone air and bone-conduction thresholds, and word recognition in quiet, is insufficient to accurately reflect the hearing loss or rehabilitation success (Binos et al., 2024). In addition to these tests, best practices in audiology suggest including measures of speech understanding in noisy environments (Billings et al., 2024), particularly for individuals who have difficulty communicating in such situations. Speech-in-Noise (SiN) testing is recognized as a better assessment of functional hearing difficulties within the World Health Organization framework (World Health Organization, 2021).

There are various SiN tests available to researchers and clinicians, including the Hearing-in-Noise Test (HINT) (Nilsson et al., 1994), the Words-in-Noise Test (WIN) (Wilson, 2003), and the Quick Speech-in-Noise Test (QuickSIN) (Killion et al., 2004). This study focuses on the QuickSIN test for the SiN assessment of hearing-impaired listeners. The QuickSIN test was chosen for investigation for the following reasons: (a) it is an efficient SiN test that is used more frequently in Audiology clinics (Billings et al., 2024; Mueller, 2016); (b) it was found to be more sensitive to individual variations in speech recognition scores and better separated two groups of listeners with normal hearing and hearing impairment (Davidson et al., 2022; Wilson et al., 2007); (c) it provides recommendations on hearing aids and assistive listening technologies based on the SiN score (Killion et al., 2004), (*OTOsuite and the QuickSIN Module User Guide*, 2019); and (d) it was found to be a significant predictor of satisfaction with hearing aids (Davidson et al., 2021).

Although the QuickSIN test is used most frequently in a clinic and a typical administration time of the QuickSIN test is approximately 4-5 minutes, it is conservatively estimated that only 10% of the audiologists routinely administer it [3], (Mueller et al., 2023). As such, a key objective of this study is to investigate the automation of QuickSIN test scoring using ASR technologies to further improve its clinical efficiency and potentially facilitate parallel testing of multiple clients.

Very few studies have employed ASR systems in automatically scoring speech-in-noise tests. Ooster et al. (Ooster et al., 2018) proposed an automated system for speech audiometry that uses automatic speech recognition (ASR) to perform the matrix sentence test for measuring the speech reception threshold (SRT). The system was evaluated using pre-recorded responses and spontaneous utterances from normal-hearing and hearing-impaired subjects, and the ASR-based SRT measurement was compared to results obtained with a human supervisor. Ooster et al. (Ooster et al., 2020) compared expert-conducted speech audiometry to self-conducted SRT measurements using smart speakers and with the matrix sentence test, finding an overall bias8 in the SRT result that depends on the hearing loss. However, the speech-based test demonstrates significant potential for complementing clinical measurements in detecting a clinically elevated SRT. The study explores differences between controlled laboratory measurements and smart speaker-based tests for young and elderly normal-hearing listeners and mildly/moderately hearing-impaired listeners across varying room acoustics (low, medium, and high reverberation). It reports an intrasubject standard deviation close to the clinical standard deviation, indicating reliability, and an area under the curve (AUC) value of 0.95 for detecting a clinically elevated SRT. The AUC, derived from the receiver operating characteristic (ROC) analysis, represents the diagnostic accuracy of the test, where a value of 1 indicates perfect discrimination and a value of 0.5 suggests no discrimination. An AUC of 0.95 reflects excellent sensitivity and specificity, signifying that the smart speaker-based SRT test effectively differentiates between clinically normal and elevated SRT levels (Ooster et al., 2020). The speech audiometry application was implemented with the Alexa Skill Developer Kit in Python and executed on an Amazon Echo loudspeaker, but it uses synthesized speech, presents sound via a speaker in a reverberant environment, stores audio files with lossy audio formats, and transcribes the listener’s response via ASR instead of being logged by an audiologist. The synthesized sentences were premixed with the speech-shaped noise at steps of 0.1 dB and then converted to the MP3 data format (MPEG version 2, 48 kbps, 16 kHz) to be played back through the smart speaker, after verifying its signal-to-noise ratio by re-recording stimuli with known SNR and analyzing them. To prevent the speaker adaptation of the ASR system, all recorded audio files in the cloud of the smart speaker were deleted at the end of each measurement session, and a human supervisor recorded the subjects’ responses during the ASR-based measurements to obtain the ground truth of responses (Abe, 2003).

In summary, only a few published studies investigated the automation of a SiN test scoring through applying ASR on the verbal responses from hearing impaired participants, with none of the studies employing the QuickSIN test. Furthermore, as highlighted in a recent scoping review on the use of ASR technologies for automating SiN testing (Fatehifar et al., 2024), there currently is a paucity of literature on the use of ASR-based SiN test scores for comparing the SiN performance associated with different hearing aid fittings. This study addresses these gaps in the literature and contributes novel information by answering the following research questions:

1. Are there significant differences between the audiologist– and ASR-derived QuickSIN scores, when the test is administered to a group of hearing-impaired listeners?
2. Are there any performance differences between contemporary ASR technologies for automatic scoring of the QuickSIN test?
3. Can ASR-based scoring of the QuickSIN test facilitate performance comparison between different hearing aid fittings?

## Methodology

### Participant and hearing aid information

Thirty-five adults with sensorineural hearing loss were recruited to participate in this study. However, during the test administration, computer instability and unforeseen issues with the custom RF protocol microphone used for recording participant verbal responses resulted in incomplete QuickSIN data from a few subjects in one or both test environments. Complete aided QuickSIN data were collected from twenty-three adults (11 females, 12 males; age range: 63-84 years, mean age: 74.4 years). Prior to the QuickSIN assessment, participants underwent a comprehensive audiometric evaluation (audiogram, tympanometry, suprathreshold tests, and real ear to coupler difference (RECD)) and cognitive screening (Montreal Cognitive Assessment (MOCA)). The average left and right ear hearing thresholds for these twenty-four adults, along with their ranges are presented in Figure 1.

**Figure 1:**
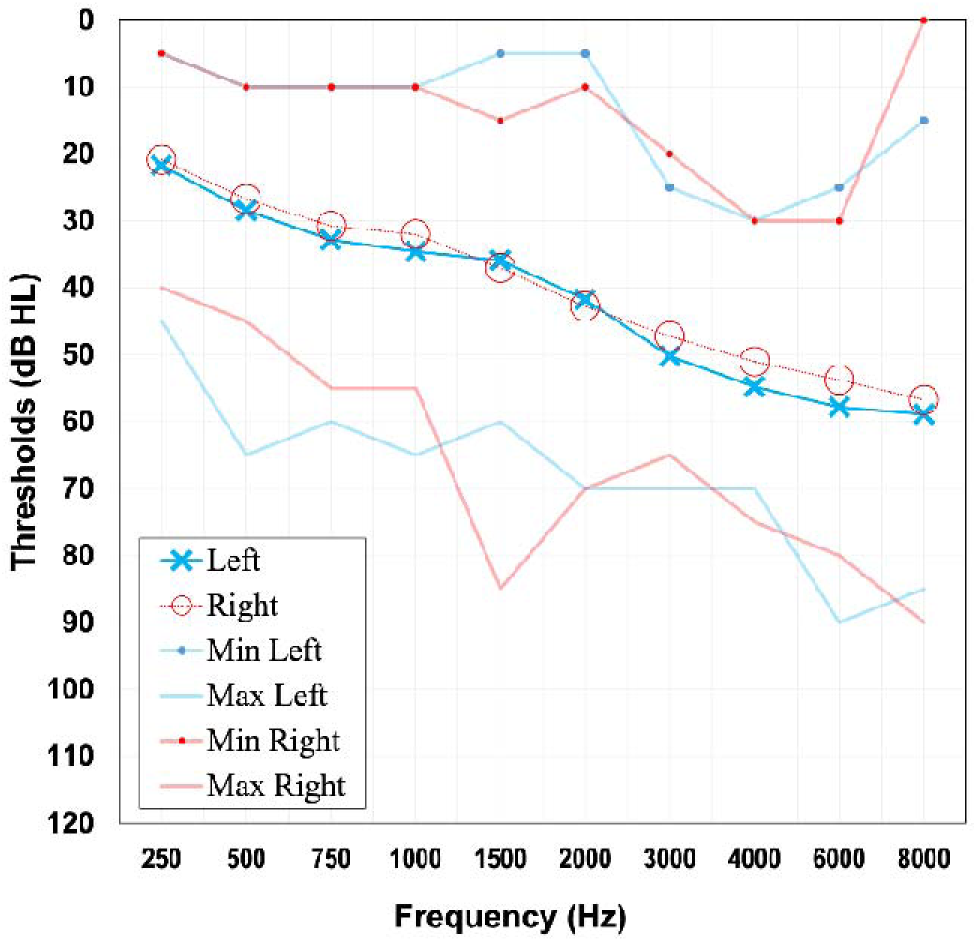
This figure shows the average hearing thresholds of the 23 participants for both left and right ears across different frequencies (0.25 – 8 kHz). In addition, the minimum and maximum threshold values at each audiometric frequency for both ears are also depicted.

All participants were fitted with bilateral Unitron D Moxi Fit 9 Receiver-in-the-canal style hearing aids with M-receivers and vented domes. The hearing aids were programmed to match the DSL v5.0 adult targets using the measured RECD values. Fit-to-targets was verified at 55-, 65-, and 75-dB SPL using the Audioscan Verifit 2. Two manual programs were created in the hearing aids – one with an omnidirectional microphone and the other with a fixed directional microphone.

All other signal processing settings such as noise reduction, speech enhancement, transient sound reduction etc. were either turned off or set to minimum strength. The feedback canceller was left at the default setting. Each participant had their own new receivers and domes that were kept in their file and before each QuickSIN appointment, the participant’s receivers and domes were placed on the hearing aids, and their custom hearing aid program settings were downloaded into the hearing aids using the manufacturer’s fitting software.

### Test conditions

The QuickSIN test was administered in two different acoustic environments: a low reverberation sound-attenuating test booth (RT_60_ = 100 ms) and a higher reverberation chamber (RT_60_ = 600 ms). While both the test environments had a circular array of loudspeakers through which speech and noise stimuli were played back, there were differences in the equipment configurations. The signal flow chain in the low reverberation environment included a multichannel soundcard (Echo AudioFire 12), programmable attenuators (Tucker Davis Technologies PA5s), multichannel speaker power amplifier (QSC CX168), and Anthony Gallo A’Diva loudspeakers. In the higher reverberation chamber, the outputs of the AudioFire multichannel soundcard were routed to a Lab Gruppen C series power amplifier, which was connected to the Tannoy dual concentric loudspeakers. In both test environments, QuickSIN speech stimuli were always played from 0° azimuth. The competing 4-talker babble was played back either from 0°, laterally, or simultaneously from 90^°^, 180^°^, and 270° azimuths, which were labeled as “0-0”, “0-90”, and “0-S” noise presentation conditions respectively. In the lateral noise condition, the babble was played to the side of the ear with poorer thresholds. If the audiograms were symmetrical, the babble was presented from the left ear side. The speech presentation level was fixed at 65 dBA, while the noise presentation level was varied based on the SNR. Test sessions in each test environment were preceded by speech and noise level calibration checks using the Larson Davis sound level meter.

### Test administration

QuickSIN administration in both test environments were facilitated through custom MATLAB software programs. Figure 2 displays the common graphical user interface (GUI) visible to the audiologist in both environments. Within the GUI, the audiologist can enter the participant and hearing aid information, select a QuickSIN list, and setup the noise presentation condition. The standard QuickSIN test is comprised of 12 lists, with each list containing 6 sentences and each sentence incorporating 5 keywords, totaling 30 keywords per list. Within the GUI, the keywords and sentences from the selected list are automatically displayed once the experiment begins. As shown towards the top-middle portion of Figure 2, a dynamic 6X 5 grid displayed the keywords for the selected list, while the entire sentences were shown below the grid. As per the QuickSIN protocol, the list sentences are presented against a background of 4-talker babble noise. The Signal-to-Noise Ratio (SNR) for these sentences began at 25 dB for the first sentence and gradually decreased to 0 dB for the final sentence. In the present study, the presentation of the background noise began immediately after clicking the Start button, but the first speech sentence was played back after a pre-programmed delay. This intentional lag was incorporated to allow sufficient settling time for the signal processing features within the hearing aids. Following the presentation of each sentence, the listener was instructed to verbally repeat what they heard. The audiologist listened to the subject’s responses through a wireless RF lavalier microphone (Alvoxcon Inc.) clipped to the subject’s shirt collar, and simultaneously pressed the corresponding keywords on the grid, where the selected keywords were highlighted in green colour.

**Figure 2:**
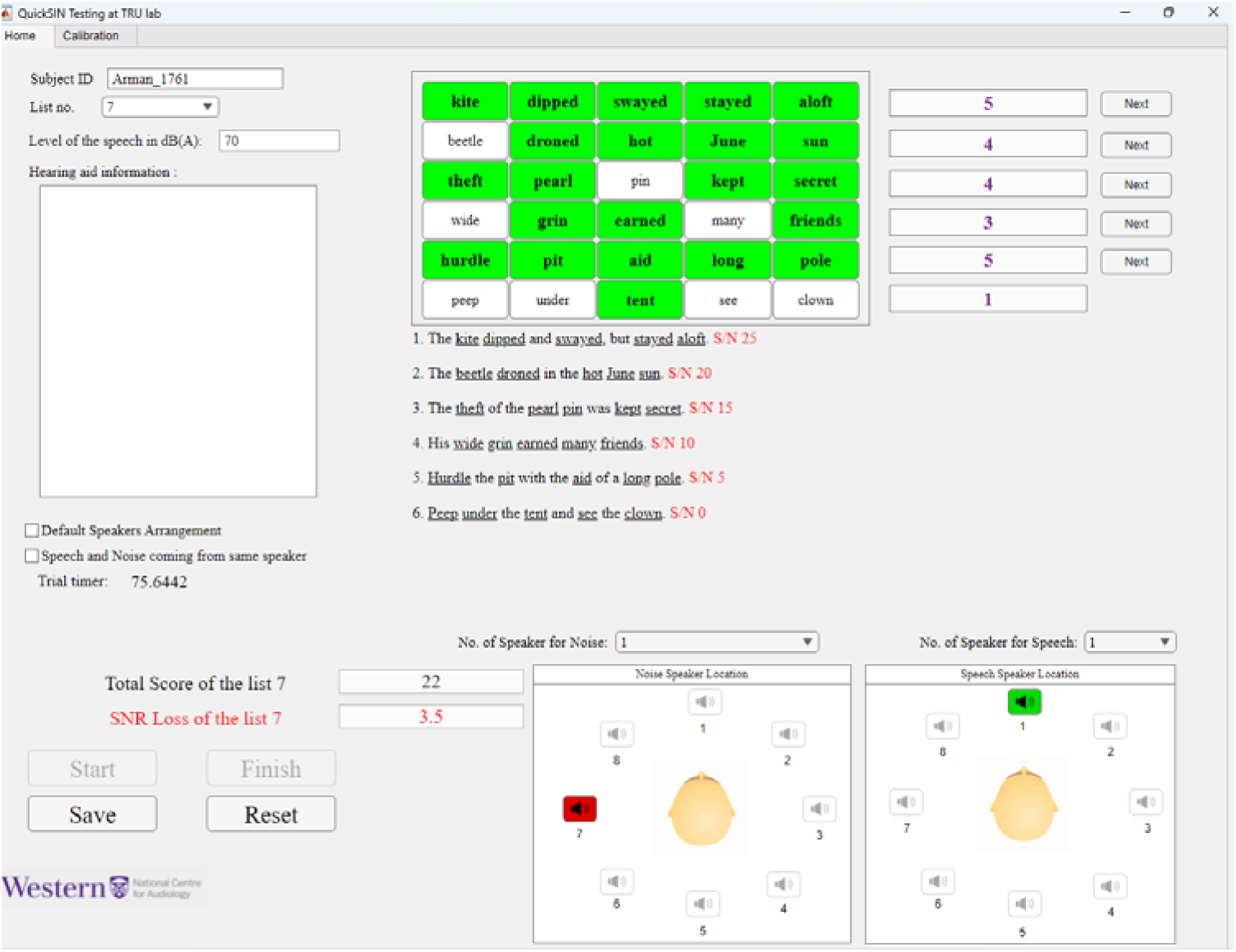
Overview of the custom QuickSIN GUI, showcasing various conditions during the presentation and recording phases. In the above chosen sound field configuration, speech is presented from 0^0^ azimuth while 4-talker babble is delivered laterally from 270^0^.

The textbox adjacent to each keypad row displayed the number of correctly scored keywords out of 5 for that sentence presentation, and clicking the Next button initiated the next sentence in the list at the appropriate SNR. At the end of the response to the last sentence, the software automatically calculated the total number of correctly identified keywords and the SNR loss. These data were automatically saved to a **.csv** file for archival and later analyses. It is pertinent to note here that the background babble continued to play in between QuickSIN sentence presentations (i.e. during the subject’s verbal response time period described below).

As an enhancement to the original QuickSIN test, the custom GUI also recorded the subject’s verbal responses. The responses were recorded in .wav format at a sample rate of 44100 Hz with 16-bit resolution and stored securely. The listener’s response, recorded between the end of the sentence presentation and the pressing of the Next sentence button, was saved with a filename that included the subject’s ID, date, and time, along with the corresponding list and background noise information (list number, spatial location, SNR etc.). These recordings were later analyzed using the ASR models, as described in the next section. As the background noise continued to persist during sentence presentations, it is pertinent to note that the verbal response recordings are contaminated with the background noise, which will increase in level throughout the test due to the sentence-by-sentence SNR decrement.

### ASR evaluation

The participants’ audio recordings were analyzed using four ASR methods: Amazon Transcribe, Microsoft Azure, NVIDIA Parakeet and the Picovoice Leopard models. The selection of Amazon Transcribe and Microsoft Azure was motivated by their commercial success and published evidence in achieving high transcription accuracy across diverse acoustic conditions. NVIDIA’s Parakeet model was included due to its state-of-the-art performance on public ASR benchmarks, where it has demonstrated high accuracy, robustness, and real time performance. To explore the feasibility of deploying ASR in resource-constrained environments, we also incorporated Picovoice Leopard, an edge-optimized ASR solution designed for low-latency, offline processing on devices such as smartphones and IoT hardware. This combination of cloud-based and edge-deployable ASR systems enables a comprehensive evaluation of both transcription accuracy and deployment flexibility in real-world scenarios. We also considered other widely used ASR systems, including those from Google, Apple, IBM, and OpenAI’s Whisper. Results revealed that Amazon, Microsoft, Whisper, NVIDIA, and Picovoice yielded comparable transcription accuracy with QuickSIN speech material (these comparative results can be found in the supplemental materials).

The Amazon, Microsoft, and NVIDIA ASR engines were desktop or cloud-based, while the Picovoice on-device ASR engine was implemented on a Smartphone. For the Amazon speech-to-text conversion, the audio files, recorded at their original sample rate of 44.1 kHz, were uploaded directly to an Amazon Web Services (AWS) S3 bucket, where they were transcribed to text (Amazon Web Services, 2024). The transcribed text was then downloaded for further analysis. Post-processing included standardizing the text by converting it to lowercase, removing punctuation, and normalizing digits. The processed text was compared to the original QuickSIN keywords to calculate metrics such as the Word Error Rate (WER) and SNR Loss. Similarly, the audio recordings at their original sample rate of 44.1 kHz were uploaded to the Microsoft Azure Speech Services, and the speech-to-text conversion commands were executed on the server. The transcribed text was then downloaded, and the same post-processing steps were applied before computing the WER and SNR Loss. For NVIDIA, the “NVIDIA Parakeet-tdt-0.6B-v2” speech recognition model was employed, which was accessed through the “NeMo” toolkit and executed in a Google Colab Pro environment with GPU acceleration (“CUDA” enabled). The transcribed text output of the NVIDIA Parakeet model was post-processed in a similar manner, and the corresponding SNR loss was calculated. Finally, the Picovoice on-device ASR engine was run through a custom iPhone app and deployed on an iPhone XR running iOS Version 17.5.1. The subjects’ verbal response recordings were transferred via AirDrop into the app’s sandbox for the speech-to-text conversion process by the Picovoice ASR model. The mobile app then selected and transcribed the recordings offline, ensuring no use of any cloud-based services. The transcribed text was displayed in a textbox within the app and subsequently imported into a Python script to tally the keywords and calculate the SNR loss. Once again, post-processing for keyword identification and QuickSIN score calculation was consistent with the cloud-based ASR models.

### Data analyses

Two QuickSIN lists were administered to each participant in each test condition and the average SNR loss across the two lists was calculated. The repeated measures ANOVA was conducted independently on the low reverberation and high reverberation room data for group-level statistical analyses. The scoring method (audiologist, Amazon, Microsoft, NVIDIA and Picovoice), the hearing aid microphone setting (omnidirectional and fixed directional), and the sound field configuration (“0-0”, “0-90”, and “0-S”) were the within-subject factors. Greenhouse-Geiser correction was used when Mauchly’s test of sphericity was violated. Post-hoc comparisons were performed on statistically significant main and interaction effects with the Bonferroni correction. In addition, Bland-Altman analyses were performed to evaluate the agreement between audiologist measurements and those obtained using the ASR models. Bland-Altman plots illustrate the differences between the methods against their average values, facilitating the assessment of whether any of the ASR algorithms can be used interchangeably with the audiologist’s method while maintaining accuracy and reliability. The plots also allow for easy visualization of the mean difference, indicating any systematic bias between the methods.

## Results

### Group-level statistical analyses

Figure 3 depicts the mean and standard deviations (as error bars) of the QuickSIN SNR loss data in the low reverberation environment, as scored by the audiologist and the four different ASR models.

**Figure 3:**
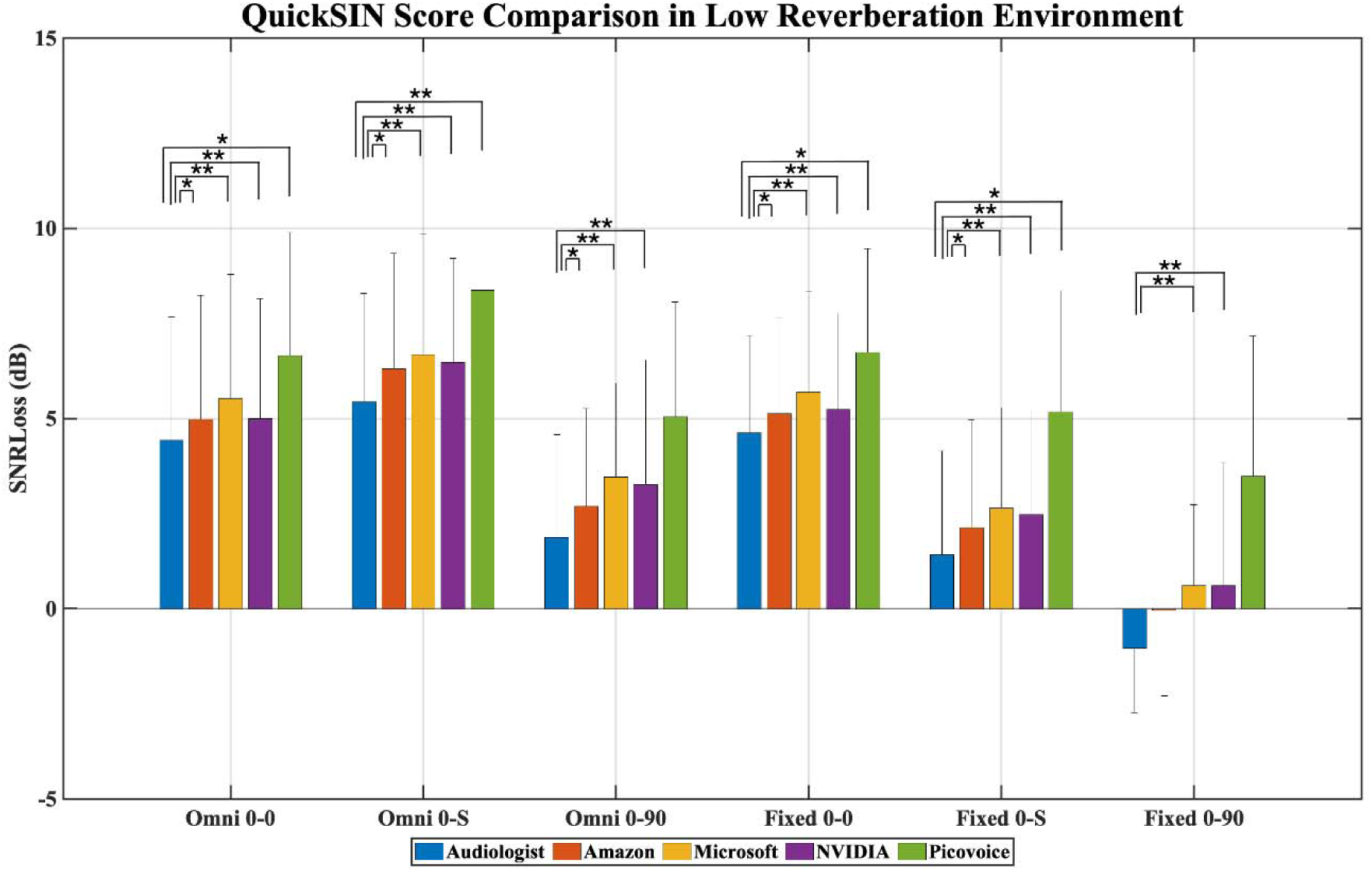
Bar plot comparing the five scoring approaches: Audiologist, and through Amazon, Microsoft, NVIDIA, and Picovoice ASR models across six different conditions in the low reverberation laboratory settings. The “∗” symbols on the bridge bars indicate significance levels associated with the multiple paired comparisons, with one “∗” for p < 0.05 and two “∗∗” for p < 0.001. In the plot, 0-0, 0-90, and 0-S denote speech presented at 0° with competing babble noise presented: 1) at 0°, 2) laterally at 90° or 270°, and 3) in a surround of sound at 90°, 180°, and 270°.

A repeated measures ANOVA revealed statistically significant main effects of both the scoring method and the measurement condition, as well as a significant interaction between the two factors, (F(4.57, 100.58) = 3.15, p = 0.013, partial η² = 0.125). On average, all ASR models produced higher SNR loss scores compared to the audiologist. As anticipated, lower SNR loss values were observed in conditions with a single lateral noise source and when the directional microphone was activated in scenarios where speech and noise were spatially separated. Pairwise comparisons with Bonferroni correction identified several statistically significant differences between scoring methods within each test condition. A complete summary of these comparisons is provided in the supplementary material. Figure 3 also addresses the primary research question by indicating the statistically significant differences between audiologist-derived and ASR-derived SNR loss scores across the various measurement conditions. Notably, the Microsoft and Picovoice ASR models consistently yielded significantly higher SNR loss values across all conditions. In contrast, the NVIDIA ASR model produced values statistically comparable to the audiologist in the “Omni 0-90” and “Fixed 0-90” conditions, while the Amazon ASR model was statistically similar to the audiologist only in the “Fixed 0-90” condition.

A similar analysis was conducted for the data collected in the high-reverberation environment. Figure 4 presents the average SNR loss values for each scoring method and measurement condition as a bar plot, with error bars representing one standard deviation across all participants for each method-condition combination. As expected, due to the presence of reverberation, SNR loss values in Figure 4 are elevated across all conditions compared to those in Figure 3.

**Figure 4:**
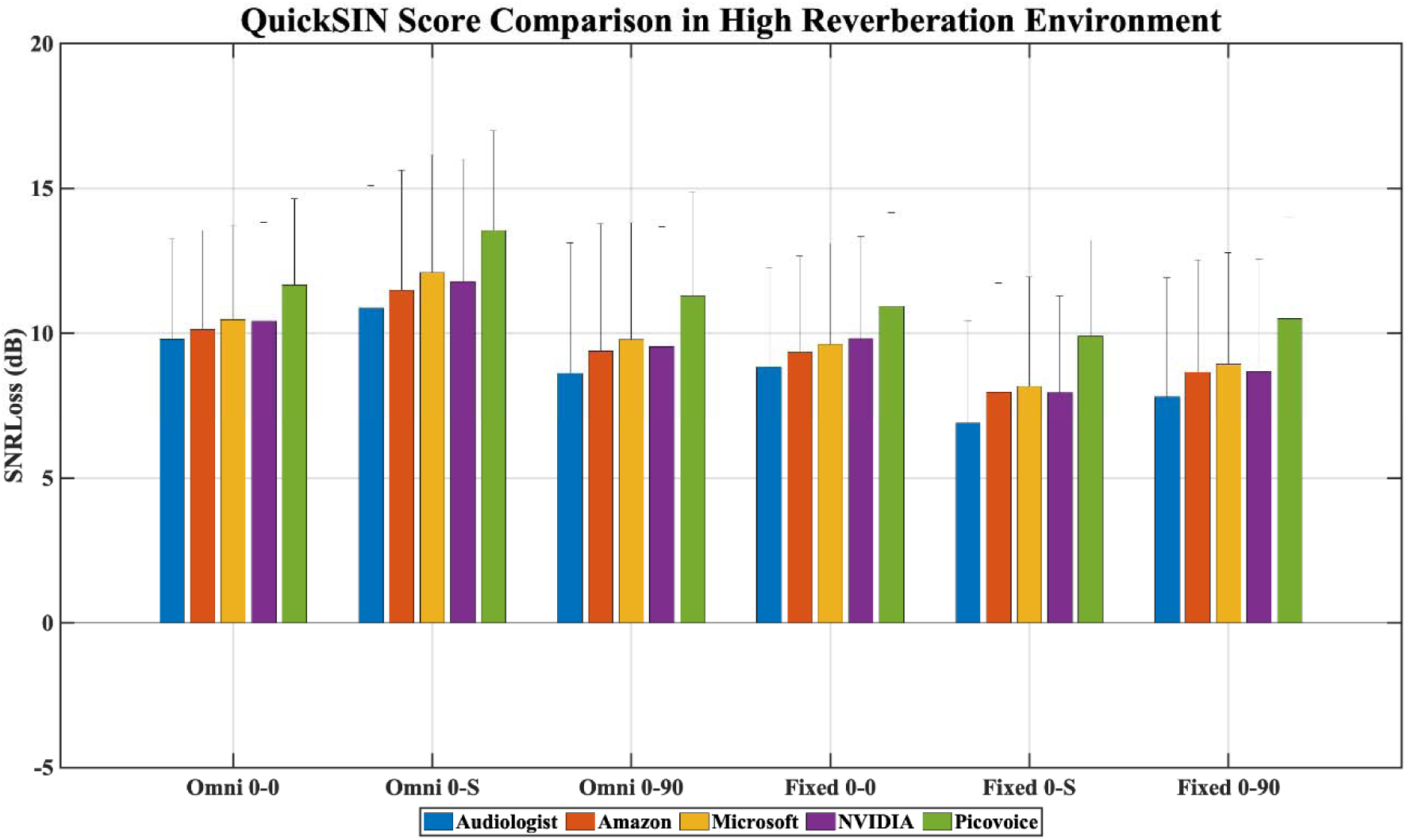
Bar plot comparing the five scoring approaches: Audiologist, and through Amazon, Microsoft, NVIDIA, and Picovoice ASR models across six different conditions in the high reverberation laboratory settings. In the plot, 0-0, 0-90, and 0-S denote speech presented at 0° with competing babble noise presented: 1) at 0^°, 2) laterally at 90° or 270°, and 3) in a surround of sound at 90°, 180°, and 270°.

Repeated measures ANOVA revealed significant main effects of scoring method (F(1.29, 235.17) = 43.25, p < 0.001, partial η² = 0.663) and measurement condition (F(5, 110) = 11.54, p < 0.001, partial η² = 0.344), with no significant interaction between the two factors (F(5.73, 254.21) = 1.24, p = 0.29). Post-hoc pairwise comparisons with Bonferroni correction revealed the following: (a) SNR loss values were highest in the “Omni 0-S” condition and significantly greater than those in all directional microphone conditions; (b) all directional microphone conditions were statistically similar to one another; (c) “Omni 0-90” scores did not differ significantly from those of the directional microphone conditions; and (d) all ASR models produced significantly higher SNR loss values than the audiologist when averaged across all measurement conditions.

Although the differences between the audiologist-derived and ASR model-derived SNR loss scores were statistically significant under certain conditions, they may not be clinically meaningful. For instance, in the low-reverberation environment, the mean absolute deviations from the audiologist scores were 0.7 dB, 1.3 dB, and 1.1 dB for the Amazon, Microsoft, and NVIDIA ASR models, respectively. These deviations fall within the 95% confidence interval of ±1.9 dB for QuickSIN scores when two lists are administered, suggesting that the discrepancies are likely within the expected range of test-retest variability. However, it is essential to examine the individual-level differences between ASR– and audiologist-derived QuickSIN scores to assess whether they consistently fall within clinically acceptable confidence bounds. This was explored through Bland-Altman analysis of the data.

### Bland-Altman analyses

Figures 5 and 6 display the Bland-Altman plots associated with the four ASR models, in the low reverberation and high reverberation environments respectively. In these figures, each subpanel depicts the scatter plot of the differences between the audiologist-derived and ASR model-derived SNR loss scores against their average, for all subjects and measurement conditions (i.e., a total of 23 subjects x 6 conditions = 138 data points in each subpanel, with test conditions denoted by different symbols). In all plots, the red dotted lines represent the upper and lower limits of agreements, while the blue solid line represents the mean difference (or mean bias) between the audiologist’s SNR loss scores and the model’s SNR loss scores across all conditions. Of particular interest in these plots are the mean bias, evidence of any systematic trend in the bias, and the extent of the limits of agreement.

**Figure 5:**
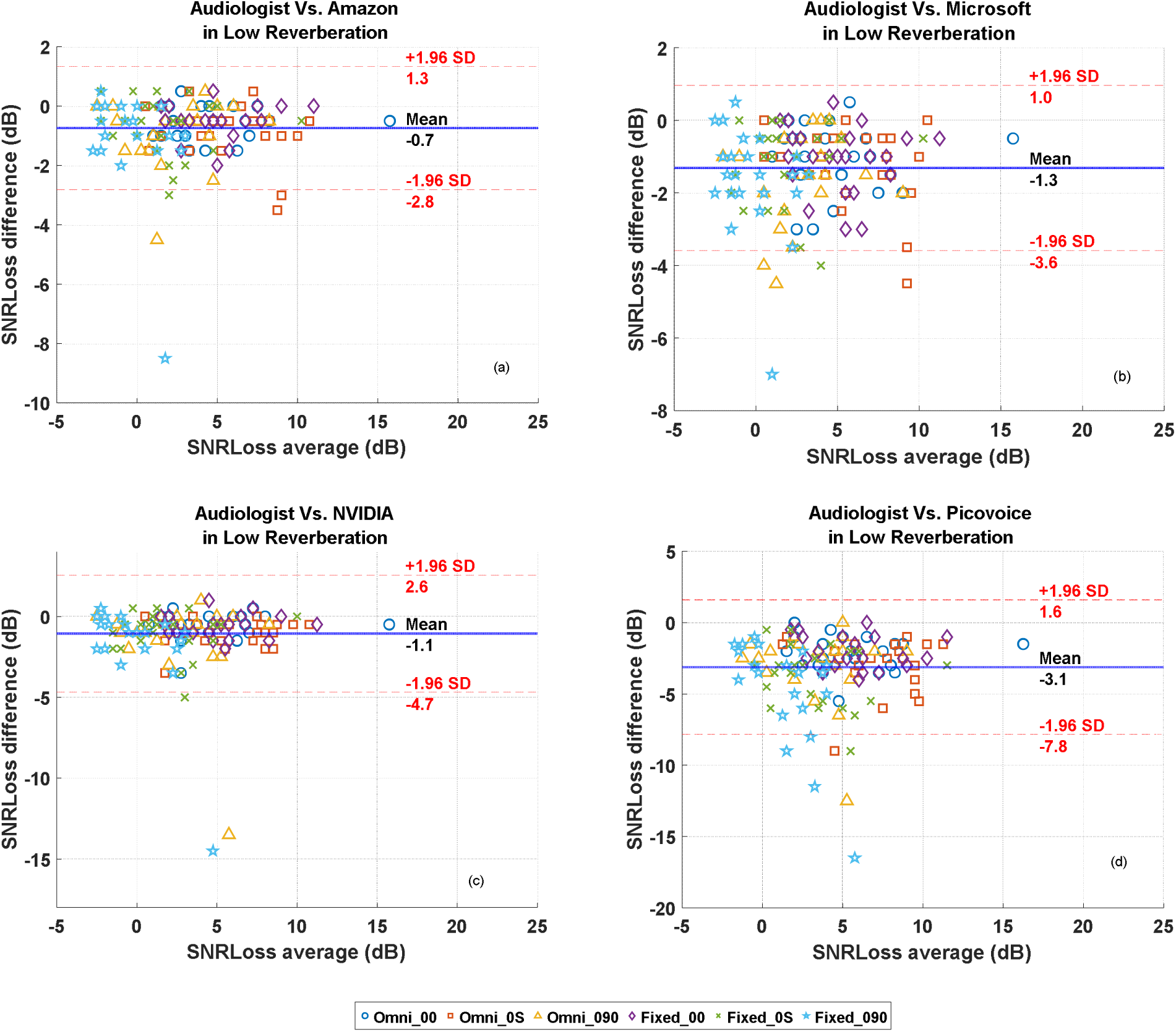
Bland-Altman plots comparing between the Audiologist– and ASR-scored SNR loss value, in the low reverberation environment. In each panel, the x-axis represents the average of the Audiologist and ASR model SNR loss values, while the y-axis represents the corresponding difference. Solid line indicates the average bias between the scoring methods, while the dashed lines indicate the limits of agreement.

**Figure 6:**
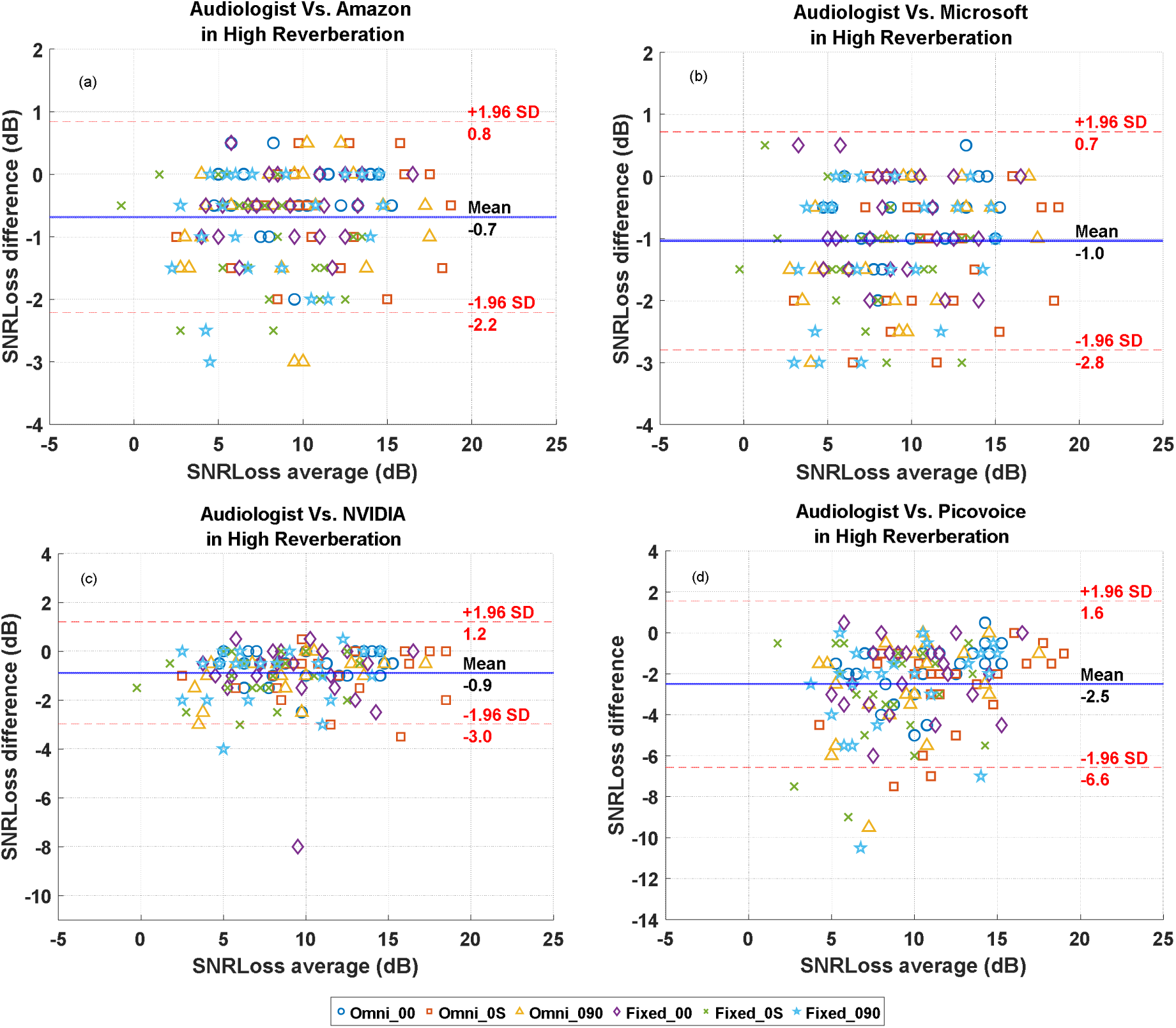
Bland-Altman plots comparing between the Audiologist– and ASR-scored SNR loss value, in the high reverberation environment. In each panel, the x-axis represents the average of the Audiologist and ASR model SNR loss values for each subject and test condition, while the y-axis represents the corresponding difference. Solid line indicates the average bias between the scoring methods, while the dashed lines indicate the limits of agreement.

As indicated in Figure 5, the mean bias for the Amazon, Microsoft, NVIDIA, and Picovoice ASR models in the low reverberation environment was –0.7 dB, –1.3 dB, –1.1 dB, and –3.1 dB, respectively. Linear regression fits to the scatter data revealed that there was no statistically significant trend across the SNR loss range for all ASR models, implying that there were no systematic differences between audiologist– and ASR model-derived SNR loss scores. The limits of agreement spanned the narrowest range for the Amazon ASR model (−2.8 dB to 1.3 dB), while they were the widest for the Picovoice ASR model (−7.8 dB to 1.6 dB).

The Bland-Altman plot associated with the high reverberation data (Figure 6) reports similar results for the mean bias and limits of agreement. The mean bias values were –0.7 dB, –1.0 dB, –0.9 dB, and –2.5 dB for the Amazon, Microsoft, NVIDIA, and Picovoice ASR models, respectively. Similar to Figure 5, the Amazon ASR model had the tightest limits of agreement (−2.2 dB to 0.8 dB), while the Picovoice ASR model had the widest (−6.6 dB to 1.6 dB), with the Microsoft (−2.8 dB to 0.7 dB) and NVIDIA (−3.0 dB to 1.2 dB) in between. Linear regression fits to the scatter data once again resulted in non-significant *F* values for Amazon, Microsoft, and NVIDIA ASR models. The data associated with the Picovoice ASR model, however, did result in a significant linear regression fit (F(1,136) = 10.82, p < 0.01). The slope of the fitted regression line was +0.16, indicating that the on-device ASR model was more deviant from the audiologist score for lower mean SNR loss values in the high reverberation environment.

### Directional benefit assessment analyses

One of the research questions pursued in this study is to investigate whether ASR-based QuickSIN scores allow for the identification of performance differences associated with different hearing aid settings. In the present study, this research question translates to whether there are any performance differences between the omnidirectional and fixed directional microphone settings across the different background noise and reverberation conditions, and whether these performance differences are captured equally by different scoring methods.

The QuickSIN guidelines (Killion et al., 2004, 2006) provide confidence intervals for comparing two different SNR loss scores from a single individual. For example, a comparison between two conditions is statistically significant at an 80% confidence interval when they differ by more than 1.8 dB for two QuickSIN lists. In this study, the difference between omnidirectional and fixed directional microphone program scores (i.e., the directional benefit) can be expected in the “0-90” and “0-S” background noise conditions in both the reverberation environments.

Table 1 displays the number of participants who have met or exceeded the threshold of 1.8 dB difference in the SNR loss scores for the directional benefit, as scored by different methods. This number is displayed for the two background noise conditions and in the two reverberation environments. Salient observations from this table include: (a) as expected more participants accrue directional benefit in the low reverberation environment than in the high reverberation environment; (b) the number of subjects meeting the criteria for critical difference was greater for the cloud-based ASR models than the on-device model; and (c) for some of the conditions, even the on-device ASR returned the same or more number of participants meeting or exceeding the threshold as the audiologist. The last point implies that both the absolute SNR loss measurement and the SNR loss difference measurement are needed for proper assessment of the ASR model performance in automatic QuickSIN scoring.

**Table 1:** Number of subjects with significant SNR loss difference* associated with directional benefit. Data is shown for different QuickSIN scoring methods and for the two reverberation environments.

| Environment | Condition | audiologist | Amazon | Microsoft | NVIDIA | Picovoice |
| --- | --- | --- | --- | --- | --- | --- |
| **Low Reverberation** | 0-S | 17 | 15 | 14 | 17 | 15 |
|  | 0-90 | 13 | 13 | 15 | 13 | 10 |
| **High Reverberation** | 0-S | 17 | 13 | 17 | 15 | 16 |
|  | 0-90 | 8 | 5 | 5 | 6 | 7 |
|  |  | *Significant SNR loss difference indicates directional benefit determined by scoring methods and reverberation environments. |  |  |  |  |

## Discussion

In the present study, the performance of modern ASR models in automatically scoring the QuickSIN test was investigated. QuickSIN scores estimated by the Amazon, Microsoft, NVIDIA, and Picovoice ASR models were compared to corresponding values manually scored by an audiologist. Both group-level statistical analyses and Bland-Altman analyses were carried out to evaluate the effectiveness of ASR-based QuickSIN scoring.

Group-level statistical analyses with repeated measures ANOVA revealed that the differences between audiologist-derived and ASR model-derived QuickSIN scores were statistically significant for most of the test conditions. Among all the ASR models, the Amazon ASR model performed the best. Statistical analyses showed that there were significant group-level differences between the audiologist and Amazon ASR model QuickSIN scores across most test settings. However, the average bias for the Amazon ASR model was only –0.7 dB in both environments, and it resulted in the tightest limits of agreement in comparison to the audiologist scores.

The negative mean values observed in the Bland-Altman plots suggest that the ASR models for automatic scoring of SIN tests tend to report higher SNR loss scores compared to those assessed by audiologists. This discrepancy likely arises because the ASR models may misinterpret or inaccurately transcribe speech, thereby attributing additional errors that are not reflective of the actual listening performance of participants. Consequently, the ASR models appear to underperform compared to audiologists, as their scoring includes transcription errors that amplify the perceived listening difficulties.

While Amazon AWS was the better performing model, there were test conditions where it (and indeed all other ASR models) deviated substantially from the audiologist score. Investigation of these test conditions revealed that the additive background noise in the subject response recordings affected the keyword recognition accuracy of the ASR models, including Amazon. As described earlier, the multi-talker babble background noise persisted in between sentence presentations during the QuickSIN test. Furthermore, the background noise level increased with each sentence presentation, as the sentence level was fixed and the SNR was varied from 25 dB to 0 dB. As such, recordings of the verbal responses to the last sentence in each list had higher background noise levels and typically lead to keyword misidentification by the ASR models. Although the audiologist listened to the same noisy verbal response, the sentence familiarity and the on-screen display of the keywords perhaps assisted in better keyword spotting during manual scoring. It is worth noting here that a single missed keyword increases the SNR loss score by 1 dB. Thus, misidentification of two keywords will place the ASR-derived score outside of the +/-1.9 dB CI for an individual QuickSIN score. As such, high quality recordings of the subjects’ responses coupled with a robust ASR model are necessary to accurately identify all the keywords repeated by the participant.

To mitigate SNR Loss accuracy issues and bring the outliers closer to the audiologist scores, we first rechecked the transcription results of the ASR models for the subject responses that led to outliers. We then examined the audio recordings for any corruption or discrepancies. As part of this process, we verified the sample rates and processed each sentence individually, rather than passing entire lists to the on-device ASR model. Some lightweight ASR models are sensitive to input length or may be trained at lower sample rates. Consequently, we downsampled all sentences from 44.1 kHz to 16 kHz for the outliers that included two lists of response recordings. However, this process did not result in any improvement in the model performance. The second approach to enhancing accuracy was the deployment of a speech enhancement (SE) algorithm as a preprocessing step before applying ASR model on the noisy recordings. However, we obtained mixed results with this particular SE algorithm – the SNR loss scores in some outlier cases, but the SE preprocessing also led to score deterioration in others.

The results associated with the resource-constrained Picovoice on-device ASR model were poorer than its more complex counterparts, suggesting that there is room for improvement. This is to be expected, as the Amazon, Microsoft, and NVIDIA models incorporate highly complex deep neural network architectures, while the Picovoice model applies a quantized version, which is more susceptible to the leakage of noise and reverberation components into the subject recording. A potential solution to enhancing an on-device model is to train a customized ASR model specifically for the QuickSIN speech stimuli, rather than deploying an ASR model trained to recognize any spontaneous speech. By optimizing hyperparameters for performance while maintaining accuracy, refining the machine learning architecture, and integrating other deep learning models, we could develop a lite version specifically for automatic QuickSIN scoring that is capable of running efficiently on edge devices.

## Limitations and future work

The manual QuickSIN scoring in this study was performed by a single audiologist. Further data collection and analyses are warranted with scoring by multiple clinical audiologists, to understand the across-audiologist scoring variability and to contrast these results with the ASR-derived data. Furthermore, data collection with a larger participant cohort size, with a more balanced distribution of participants in different hearing loss categories is warranted to further quantify the performance of the ASR models in automatically scoring the QuickSIN. In a similar vein, further data collected with different hearing aid models and signal processing settings will allow for drawing more generalizable conclusions on ASR-based QuickSIN scoring.

For optimal ASR accuracy, proper gating and isolation of the listener’s verbal responses to QuickSIN stimuli were found to be crucial. Additionally, accurate transcription heavily depends on the correct positioning of the recording microphone to minimize the impact of background noise. In future studies, utilizing beamforming and noise-cancelling microphones may further help mitigate the effects of background noise and reverberation in ASR-based automatic QuickSIN scoring.

While the present study provides valuable information on how close the ASR-derived QuickSIN scores are to audiologist scores, further work is necessary to directly quantify the 95% CI for QuickSIN scores when ASR methods are used. This applies to both establishing the reliability of an individual ASR-computed QuickSIN score, and for comparing ASR-derived QuickSIN scores across two conditions. Such an investigation would require the estimation of standard deviations associated with the test-retest and across-list QuickSIN scores obtained from a group of hearing-impaired listeners using the chosen ASR-based method.

## Data Availability

Data Availability Statement: All data produced and analyzed in the present study including QuickSIN scores, ASR transcriptions, and related statistical analyses are available upon reasonable request to the corresponding author. Due to ethical constraints associated with participant privacy and institutional research ethics board (REB) approval, raw audio recordings are not publicly shared but may be made available under appropriate data sharing agreements. Supplementary performance metrics and tables are included in the manuscript and supplementary material.

## Acknowledgements

The authors would like to thank Steve Beaulac and Matt Holden from the National Centre for Audiology, and Leonard Cornelisse from Unitron Canada for their technical support.

## Funding

This research was supported by the Ontario Research Fund Grant RE08-072 (PI: Dr. Susan Scollie) and the NSERC Discovery Grant to Dr. Vijay Parsa.

## Conflicts of Interest

No conflicts of interest are reported.

## Supplementary Information

**Table S1:** Comparison of ASR model performance with a set of QuickSIN lists.

|  | List 1 |  | List 2 |  | List 6 |  | List 8 |  | List 10 |  | List 11 |  | List 12 |  | List 1, 2, 6, 8,10,11,12 |
| --- | --- | --- | --- | --- | --- | --- | --- | --- | --- | --- | --- | --- | --- | --- | --- |
|  | % Correct | Total Correct (Out of 30) | % Correct | Total Correct (Out of 30) | % Correct | Total Correct (Out of 30) | % Correct | Total Correct (Out of 30) | % Correct | Total Correct (Out of 30) | % Correct | Total Correct (Out of 30) | % Correct | Total Correct (Out of 30) | Average % Correct |
| Google Cloud | 90 | 27 | 90 | 27 | 93 | 28 | 83 | 25 | 77 | 23 | 70 | 21 | 97 | 29 | 180/210=<br>**0.86** |
| Microsoft Azure | 97 | 29 | 97 | 29 | 97 | 29 | 83 | 28 | 93 | 28 | 93 | 28 | 100 | 30 | 201/210=<br>**0.96** |
| Amazon Web Service | 97 | 29 | 100 | 30 | 100 | 30 | 97 | 29 | 97 | 29 | 97 | 29 | 100 | 30 | 206/210=<br>**0.98** |
| IBM Cloud | 93 | 28 | 100 | 30 | 87 | 26 | 90 | 27 | 93 | 28 | 87 | 26 | 93 | 28 | 193/210=<br>**0.92** |
| Google | 80 | 24 | 90 | 27 | 93 | 28 | 83 | 25 | 93 | 28 | 63 | 19 | 80 | 24 | 175/210=<br>**0.83** |
| Houndify | 93 | 28 | 93 | 28 | 87 | 26 | 97 | 29 | 100 | 30 | 93 | 28 | 97 | 29 | 198/210=<br>**0.94** |
| DeepSpeech Mozilla | 87 | 26 | 77 | 23 | 87 | 26 | 83 | 28 | 90 | 27 | 93 | 28 | 97 | 29 | 187/210=<br>**0.89** |
| WitAI | 73 | 22 | 73 | 22 | 77 | 23 | 73 | 22 | 77 | 23 | 63 | 19 | 83 | 25 | 156/210=<br>**0.74** |
| Voice Overlay | 87 | 26 | 97 | 29 | 90 | 27 | 77 | 23 | 80 | 24 | 83 | 25 | 93 | 28 | 182/210=<br>**0.87** |
| Leopard | 97 | 29 | 97 | 29 | 97 | 29 | 90 | 27 | 100 | 30 | 90 | 27 | 97 | 29 | 200/210=<br>**0.95** |
| NVIDIA Parakeet | 100 | 30 | 97 | 29 | 100 | 30 | 97 | 29 | 97 | 29 | 100 | 30 | 100 | 30 | 207/210=<br>**0.99** |

## Notes

### Competing Interest Statement

The authors have declared no competing interest.

### Author Declarations

The Health Sciences Research Ethics Board (HSREB) of Western University gave ethical approval for this work. Approval was issued on April 23, 2024, for the study titled 'Speech in Noise Test Scoring Using Automatic Speech Recognition', Project ID 124196, Review Reference 2024-124196-91975.

## References

1. Abe, S. (2003). Analysis of Multiclass Support Vector Machines. Proc. International Conference on Computational Intelligence for Modelling Control and Automation (CIMCA’2003), 385–396.

2. Amazon Web Services. (2024). Security and Privacy in Amazon Transcribe. https://docs.aws.amazon.com/transcribe/latest/dg/security.html

3. Billings, C. J., Olsen, T. M., Charney, L., Madsen, B. M., & Holmes, C. E. (2024). Speech-in-Noise Testing: An Introduction for Audiologists. Seminars in Hearing, 45(01), 055–082. 10.1055/s-0043-1770155

4. Binos, P., Korres, G., Papastefanou, T., Papadimitriou, N., & Psillas, G. (2024). From Pure Tones to Complex Sounds: Expanding Audiology Tools to Better Address Speech and Language Development. Cureus, 16(10), e72519.

5. Davidson, A., Marrone, N., & Souza, P. (2022). Hearing Aid Technology Settings and Speech-in-Noise Difficulties. American Journal of Audiology, 31(1), 21–31. 10.1044/2021_AJA-21-00176

6. Davidson, A., Marrone, N., Wong, B., & Musiek, F. (2021). Predicting Hearing Aid Satisfaction in Adults: A Systematic Review of Speech-in-noise Tests and Other Behavioral Measures. Ear & Hearing, 42(6), 1485–1498. 10.1097/AUD.0000000000001051

7. Fatehifar, M., Schlittenlacher, J., Almufarrij, I., Wong, D., Cootes, T., & Munro, K. J. (2024). Applications of automatic speech recognition and text-to-speech technologies for hearing assessment: A scoping review. International Journal of Audiology, 1–12. 10.1080/14992027.2024.2422390

8. Gulati, A., Qin, J., Chiu, C.-C., Parmar, N., Zhang, Y., Yu, J., Han, W., Wang, S., Zhang, Z., Wu, Y., & Pang, R. (2020). Conformer: Convolution-augmented Transformer for Speech Recognition (No. arXiv:2005.08100). arXiv. http://arxiv.org/abs/2005.08100

9. Killion, M. C., Niquette, P. A., & Gudmundsen, G. I. (2004). Development of a quick speech-in-noise test for measuring signal-to-noise ratio loss in normal-hearing and hearing-impaired listenersa). The Journal of the Acoustical Society of America, 116(4), 2395–2405.

10. Killion, M. C., Niquette, P. A., Gudmundsen, G. I., Revit, L. J., & Banerjee, S. (2006). Erratum:“Development of a quick speech-in-noise test for measuring signal-to-noise ratio loss in normal-hearing and hearing-impaired listeners”[J. Acoust. Soc. Am. 116 (4), 2395–2405 (2004)]. The Journal of the Acoustical Society of America, 119(3), 1888–1888.

11. Mueller, H. G. (2016, September). Signia expert series: Speech-in-noise testing for selection and fitting of hearing aids: Worth the effort. AudiologyOnline. https://www.audiologyonline.com

12. Mueller, H. G., Ricketts, T., & W. Y. Hornsby, B. (2023, November 6). 20Q: Speech-in-Noise Testing—Too Useful to be Ignored! AudiologyOnline. https://www.audiologyonline.com/articles/20q-speech-in-noise-testing-28760

13. Nilsson, M., Soli, S., & Sullivan, J. (1994). Development of the Hearing In Noise Test for the measurement of speech reception thresholds in quiet and in noise. 95(2), 1085–1099.

14. Ooster, J., Huber, R., Kollmeier, B., & Meyer, B. T. (2018). Evaluation of an automated speech-controlled listening test with spontaneous and read responses. Speech Communication, 98, 85–94. 10.1016/j.specom.2018.01.005

15. Ooster, J., Krueger, M., Bach, J.-H., Wagener, K. C., Kollmeier, B., & Meyer, B. T. (2020). Speech Audiometry at Home: Automated Listening Tests via Smart Speakers With Normal-Hearing and Hearing-Impaired Listeners. Trends in Hearing, 24, 233121652097001. 10.1177/2331216520970011

16. *OTOsuite and the QuickSIN Module User Guide*. (2019). https://partners.natus.com/asset/resource/file/otometrics/asset/2019-07/7-50-1200-EN_03.PDF

17. Shin, W., Lee, B. H., Kim, J. S., Park, H. J., & Han, S. W. (n.d.). MetricGAN-OKD: Multi-Metric Optimization of MetricGAN via Online Knowledge Distillation for Speech Enhancement.

18. Wilson, R. H. (2003). Development of a Speech-in-Multitalker-Babble Paradigm to Assess Word-Recognition Performance. Journal of the American Academy of Audiology, 14(9), 453–470.

19. Wilson, R. H., McArdle, R. A., & Smith, S. L. (2007). An Evaluation of the BKB-SIN, HINT, QuickSIN, and WIN Materials on Listeners With Normal Hearing and Listeners With Hearing Loss. Journal of Speech, Language, and Hearing Research, 50(4), 844–856. 10.1044/1092-4388(2007/059)

20. World Health Organization. (2021). World Report on Hearing. World Health Organization. https://www.who.int/publications/i/item/world-report-on-hearing

